# TENDINopathy Severity assessment – Achilles (TENDINS-A): Evaluation of reliability and validity in accordance with COSMIN recommendations

**DOI:** 10.1101/2023.10.05.23296581

**Authors:** Myles C Murphy, Fergus McCleary, Dana Hince, Ruth L Chimenti, Paola T Chivers, J. Turner Vosseller, Sophia Nimphius, Nonhlanhla Mkumbuzi, Peter Malliaras, Nicola Maffuli, Robert-Jan De Vos, Ebonie Rio

## Abstract

**Objectives:** Evaluate the construct validity (structural validity and hypothesis-testing), reliability (test-retest reliability, measurement error and internal consistency) and minimal detectable change (MDC) of the 13-item TENDINopathy Severity assessment–Achilles (TENDINS-A).

**Methods:** Participants with Achilles pain completed an online survey: demographics, TENDINS-A, Foot and Ankle Outcome Score (FAOS) and Victorian Institute of Sport Assessment-Achilles (VISA-A). Exploratory factor analysis (EFA) assessed dimensionality. Confirmatory Factor Analysis (CFA) assessed structural validity [root-mean-squared error of approximation (RMSEA); Comparative Fit Index (CFI); Tucker-Lewis Index (TLI); Standardised Root Measure Square (SRMS)]. Correlations between TENDINS-A and the FAOS/ VISA-A assessed hypothesis-testing. Intraclass correlation (ICC) assessed test-retest reliability. Cronbach’s α assessed internal consistency. Standard error of the measurement (SEM) assessed measurement error. A distribution-based approach assessed MDC.

**Results:** Seventy-nine participants (51% female) with a mean (SD) age=42.6 (13.0)years, height=175.0 (11.7)cm and body mass=82.0 (19.1)kg were included. EFA identified three meaningful factors, proposed to be pain, symptoms and function. The best model identified using CFA had adequate structural validity (CFI= 0.959, TLI= 0.947, RMSEA= 0.101, SRMS=0.068) excluded three items from the original TENDINS-A, included three factors (Pain, Symptoms, and Function). TENDINS-A exhibited moderate positive correlation with FAOS (rho=0.598,p<0.001), moderate, negative correlation with VISA-A (r=-0.639,p<0.001). Reliability of the TENDINS-A is excellent (ICC=0.930; Cronbach’s α=0.808; SEM=6.54 units) and has an MDC of 12 units.

**Conclusions:** Our evaluation of the revised 10-item TENDINS-A has determined it has adequate validity and reliability. Thus, the TENDINS-A can be recommended for immediate use, being the preferred tool over other PROMs to assess disability in Achilles tendinopathy.

## INTRODUCTION

Achilles tendinopathy is characterised by focal Achilles tendon pain accompanied by impaired function with mechanical loading,^1^ which is typically managed using exercise rehabilitation.^2 3^ Achilles tendinopathy is common in the running population^4^ and also prevalent within the general population.^5^ People with tendinopathy, expert clinicians and researchers have identified several core health domains as important in tendinopathy, including tendon-related disability.^6^ However, multiple different measures to quantify tendon-related disability are used across studies, with no existing consensus.^7 8^ In clinical practice measures of tendon-related disability are not commonly used,^9^ in part due to inadequate content validity.^10 11^ Thus, despite a consensus of it being an important aspect of health,^6^ there remains no measure of tendon-related disability that is validated for research or clinical use.

The consensus-based standards for the selection of measurement instruments (COSMIN) guidelines represent international best-practice for the design and appraisal of Patient Reported Outcome Measures (PROMs).^12 13^ However, the value of a PROM is not in its availability but in its quality, reflected by its clinometric properties.^14^ Valid and reliable PROMs should be used and recommended, but this is not currently possible in Achilles tendinopathy research of the disability domain.^15^ Existing PROMs, such as the Victorian Institute of Sport Assessment – Achilles (VISA-A), are recommended in the absence of other tools,^16^ but do not satisfy the guidelines set out by COSMIN for the assessment of Achilles tendinopathy, largely because of inadequate content and structural validity.^10 11^

The newly developed TENDINopathy Severity assessment – Achilles (TENDINS-A) has adequate content validity, being co-designed by people with Achilles tendinopathy, expert clinicians and researchers, to assess ‘disability’ from Achilles tendinopathy.^17^ TENDINS-A consists of questions covering sub-domains of pain, symptoms, and function related to Achilles tendinopathy.^17^ Whilst the TENDINS-A has sufficient content validity, the structural validity and other measurement properties of this new PROM (e.g., reliability) remain unknown.

### Objective

The objective of this study was to evaluate the construct validity (including structural validity and hypothesis testing), reliability (including test-retest reliability, measurement error and internal consistency) and minimal detectable change of the TENDINS-A.

### Hypotheses

The TENDINS-A will have adequate criteria-based structural validity, demonstrate moderate significant correlations to the VISA-A and Foot and Ankle Outcome Score (FAOS), no correlation to unrelated constructs such as baseline characteristics (e.g., age or body mass index) and excellent reliability.

## METHODS

### Study Design

Cross-sectional cohort study evaluating the psychometric properties of the TENDINS-A.

### Participants and setting

We used a network of clinicians (exercise scientists, general practitioners, orthopaedic surgeons, physiotherapists, podiatrists, rheumatologists and sport and exercise physicians) and Achilles tendon researchers to identify participants with Achilles tendinopathy and provide them with our online survey using Qualtrics. This was accessed via a quick response (QR) code or anonymous web link. Any person over the age of 18 years, who could read the English language, with self-reported Achilles tendinopathy was eligible.

### Inclusion criteria

Participants were provided with a pain map with established locations of Achilles tendon symptoms and asked to select all markings that corresponded to their region of pain,^18^ which is an accepted method to identify patients with Achilles tendinopathy.^19^

### Outcome measures

All outcome measures were completed by participants within a single survey and the survey forced responses to avoid missing data.

#### Participant characteristics

Age (years), sex (male/female/intersex), height (cm), body mass (kg), ethnicity, country of residence, languages other than English spoken by the participant (self-reported), whether the participant performed moderate to vigorous physical activity (MVPA) most days (yes/no), highest level of education, work status and total household income were reported.

#### TENDINopathy Severity assessment – Achilles (TENDINS-A)

The 13-item TENDINS-A was provided to participants as the first PROM within the survey and was scored between 0 and 100, with ‘0’ representing a perfect score (no disability) and ‘100’ representing complete disability.^17^ If participants were unable to perform one of the pain-with-loading tests (e.g., single leg hop), they were instructed to leave it blank and a score of ‘10’ was provided.

Participants completed the TENDINS-A prior to the FAOS and the VISA-A. Directly after completing the VISA-A questionnaire, the TENDINS-A was immediately repeated. This ensured the clinical status of participants was unchanged,^20^ whilst still allowing for any potential interference effects. Specifically, the participant would be unable to remember responses to the initial TENDINS-A as they have no option to go back in the survey and 50 questions (i.e., the VISA-A and FAOS) separated the initial and then repeated TENDINS-A.

#### Foot and Ankle Outcome Score

The 42-item FAOS^21^ was performed following the TENDINS-A, with a mean total score calculated for the purposes of this study. A score of ‘0’ represented a perfect score (no disability), with a score of ‘100’ representing complete disability.

#### Victorian Institute of Sport Assessment – Achilles

The 8-item VISA-A^22^ was performed following the FAOS. The VISA-A is inversely scored when compared to the TENDINS-A and FAOS with a score of ‘100’ represented a perfect score (no disability), and a score of ‘0’ representing complete disability. Where participants selected multiple responses in item 8 (related to how much physical activity can be performed before cessation due to symptoms is required), the lowest score was retained for analysis.

### Power calculation

Study size was informed by the recommendations of the COSMIN risk of bias checklist^12 13^ rather than formal power calculations. Questions 1, 2, 5 and 9 are not scored in the TENDINS-A; therefore the PROM was initially considered to have 13 scorable items as one item has five secondary scales.^17^ COSMIN guidelines suggest that six-times the number of persons to items is adequate for assessing structural validity using classical test theory,^12 13^ thus the minimum sample size for this study was determined to be 78 persons. We also required all communalities in exploratory factor analysis (EFA) to be >0.3, thus samples of <100 participants are justified.^23^ Furthermore, a sample of >50 participants is considered adequate for reliability and internal consistency.^12 13^

### Statistical analysis

The different statistical analyses, per each measurement property, are described below. All statistical analyses were performed within IBM SPSS statistics (version 29.0) or IBM SPSS Amos (version 29.0). Where appropriate, confidence intervals are presented, and statistical significance was considered when p<0.05.

#### Dimensionality

We performed EFA for all scoreable items using SPSS Statistics to assess dimensionality. To proceed to confirmatory factor analysis (CFA), we required all communalities to exceed 0.3,^24^ required the Kaiser-Meyer-Olkin Measure of Sampling Adequacy to exceed 0.7^25^ and for Bartlett’s test of Sphericity to be significant (p<0.01).^26^

Factors were eligible for inclusion within EFA where the Eigenvalues exceeded 1.0. The Scree plot was also visually inspected and additional models were performed to include more/ less factors, based on author (MCM) judgement and with the TENDINS-A being proposed to measure three factors related to disability: pain, symptoms and function.^17^ A principal axis factoring method was used to determine our initial factor matrix.

Factors were considered meaningful when an item loaded at ≥0.4 for that factor.^27^ As ‘disability’ is a domain that reflects pain, symptoms and function,^6^ we hypothesised the TENDINS-A to have three factors identified by EFA.

#### Structural Validity

Confirmatory factor analysis was performed using SPSS Amos (22) to assess structural validity. We included all three factors (which were identified in the EFA and TENDINS-A content validity study),^17^ within the initial model, and then removed variables as needed to achieve the best fit. Items with categorical (yes/ no only) responses were not included within CFA. Models were tested and selected based off the best root-mean-squared error of the approximation (RMSEA) and overall model chi-squared statistics, with a lower chi-squared statistic representing a better model fit. An overall acceptable model would require a Comparative Fit Index (CFI) OR Tucker-Lewis Index (TLI) of >0.95 AND RMSEA of <0.06 or Standardised Root Measure Square (SRMS) of <0.08.^12^

The CFA provided standardised factor loading and error variance per item. Composite reliability of >0.70 is considered adequate.^28^ Composite reliability was calculated using the formula below when λ = standardised factor loading and ε = error variance:

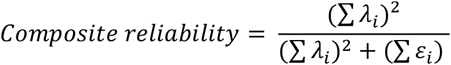

Convergent validity was assessed using the average variance extracted (AVE). An AVE of >0.50 is considered adequate.^29^ The AVE was calculated using the formula below when λ = standardised factor loading and ε = error variance:

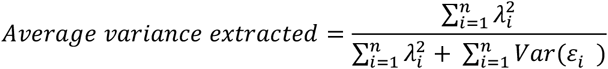

The criteria for adequate structural validity of the final model were:

1. CFI or TLI >0.95,^12^
2. RMSEA of <0.06 or SRMS of <0.08,^12^
3. Composite Reliability >0.70,^28^ and
4. 4. AVE >0.50.^29^

#### Test-retest reliability

Test-retest reliability of the TENDINS-A was calculated as a continuous scale. The absolute and relative test-retest reliability (which is a sub-type of reliability and demonstrates how closely someone will generate the same score on a PROM when repeated in a specified timeframe) were determined.^30^ Relative test-retest reliability was reported as the intraclass correlation (ICC), with a two-way mixed approach used to assess absolute agreement of a single measure.^31^ Absolute test-retest reliability is a sub-type of reliability and demonstrates the degree of uncertainty in a measurement. For example, measurements with more uncertainty will have greater measurement error, hence it is systematic and random error that is not reflective of true change.^31^ The standard error of the measurement^32^ was calculated using the following equation:

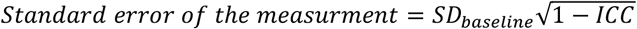

#### Minimal detectable change

The Minimal detectable change (MDC) and 95% confidence intervals was calculated using a distribution-based method,^33^ using the following equation:

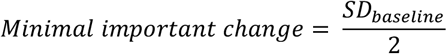

#### Internal consistency

Internal consistency is a sub-type of reliability that investigates whether different items that measure the same construct give comparable outcomes. The internal consistency for the entire 13-item TENDINS-A was calculated using the average inter-item correlation and a Cronbach’s Alpha was reported with a positive rating given for values >0.7.^12^

#### Convergent and divergent validity

In addition to the convergent validity assessed using CFA, we also assessed convergent and divergent validity of the TENDINS-A scale using a correlation between scores of the TENDINS-A and another PROM theorised to measure the same overall core health domain of tendinopathy (disability)^6 15^ or unrelated constructs, such as age and BMI (given they are continuous variables that should not be related to tendon-related disability). The TENDINS-A, FAOS and the VISA-A were assessed for normality using visual assessment and the one-sample Kolomogorov-Smirnov Test. For normally distributed data the Pearson’s correlation coefficient was used and for non-normally distributed data the non-parametric Spearman rho correlation was used. Correlations were considered very weak (between 0-0.25), weak (0.26-0.49), moderate (0.5-0.69), strong (0.7-0.89) and verystrong (0.9-1).

#### Comparison between sub-groups

Between-group differences for key baseline characteristics [sex (male/ female), MVPA most days (yes/no), multilingual status (yes/ no), and whether tendon loading activities are typically performed daily (yes/no)] were calculated using independent t-tests.

## RESULTS

### Participants

Seventy-nine (n=79) participants (51% female, 49% male) provided survey data and were included within our analysis with no missing data. Participants had a mean (SD) age of 42.6 (13.0) years, height of 175.0 (11.7) cm and body mass of 82.0 (19.1) kg. Participants were predominantly of ‘Australian’ ethnicity (79.7%) and were not multi-lingual (64.6%) with those multilingual participants predominantly having English as their first language (53.6%). A significant number of participants reported that they performed MVPA most days (69.6%). Participant education level and employment varied, with most having completed tertiary studies (74.6%) and working full time (63.3%). Income levels ranged from less than $30,000 Australian Dollars (AUD) (5.1%) to greater than $200,000 AUD (27.8%), per annum. The activities that aggravated the pain of participants with Achilles tendinopathy varied: walking slow (15.2%), walking fast (44.3%), walking up and down stairs (43.0%), running up and down stairs (46.8%), running slow (60.8%), running fast (54.4%), hopping and jumping (62.0%), and rapidly changing direction while running (44.3%). The complete breakdown of participant characteristics is provided in Appendix A.

### Uni-dimensionality

Bartlett’s test of Sphericity suggested that the results of our correlation matrix were not random [χ^2^(78) = 708, p<0.001]. Our Kaiser-Meyer-Olkin Measure of Sampling Adequacy (0.811) far exceeded the minimum cut-off of 0.70. Thus, our correlation matrix was appropriate. The extracted communalities (Mean=0.67, SD= 0.16, Range= 0.18 to 0.93) for all but three items (items 6, 8 and 12) exceeded our cut-off score for CFA inclusion (Appendix B).

Principal axis factoring analysis was performed and the initial Eigen values of three factors exceeded 1.0 (see appendix C), explained 67.9% of the total variance and the first factor accounted for >20% of the variability. The three identified factors were included within the Factor Matrix, and associated factor loading per item, are presented within Table 1. However, no items were associated with a loading of >0.4 on Factor Three.

**Table 1.** Factor Loading with Principal Axis Factoring Analysis for all items with loading >0.4 (n=79).

| TENDINS- A | Factor One | Factor Two | Factor Three |
| --- | --- | --- | --- |
| Item 3 | 0.412 |  | 0.400 |
| Item 4 | 0.496 | 0.479 |  |
| Item 6 |  |  |  |
| Item 7 |  | 0.644 |  |
| Item 8 |  | 0.504 |  |
| Item 10 |  | 0.672 |  |
| Item 11 | 0.614 |  |  |
| Item 12 |  |  |  |
| Item 13A | 0.731 |  |  |
| Item 13B | 0.888 |  |  |
| Item 13C | 0.915 |  |  |
| Item 13D | 0.916 |  |  |
| Item 13E | 0.917 |  |  |

\*blank cells represent loading of <0.4

### Structural Validity

Several Maximum Likelihood Estimates CFA models were trialled, informed by the factor loading from EFA, and subscales from our content validity study to generate our final CFA model. The best model used three factors [a) Pain, b) Symptoms, and c) Function] and eight items (Figure 1). Question 12 was excluded because of poor EFA initial communality, non-meaningful factor loading within EFA and being unable to load onto either of the three factors within CFA. Question 13A and 13B were removed as their exclusion resulted in substantially improved model fit (Appendix D). The scoring for the remaining 12 questions was amended to maintain a score range from 0 to 100, as presented within Appendix E.

**Figure 1.**
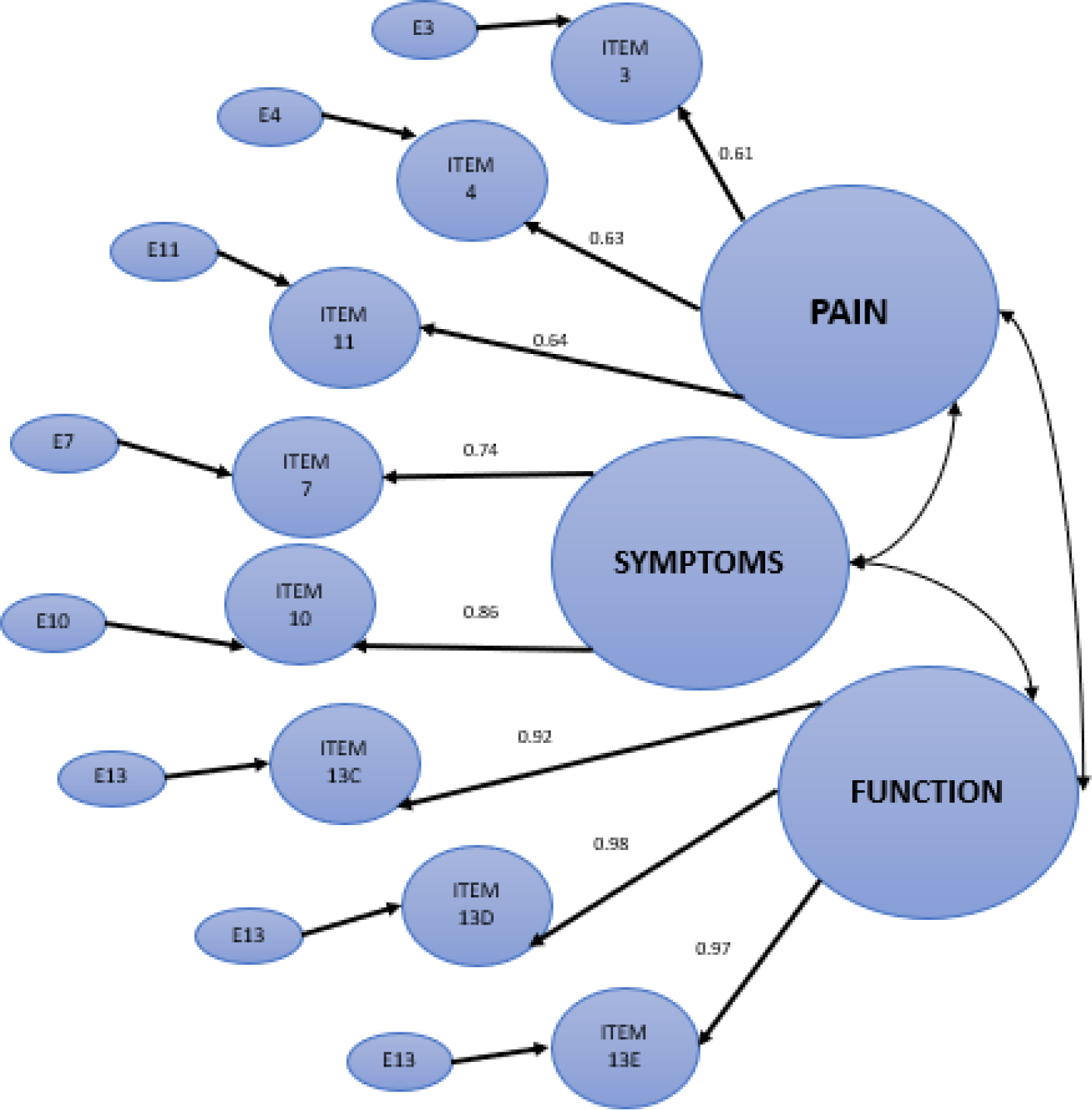
Factor Loading within Confirmatory Factor Analysis.

In the final model, the chi-squared statistic was χ^2^(22)= 39.2, the CFI was 0.959, the TLI was 0.947, the RMSEA was 0.101 and the SRMS was 0.068, indicating sufficient structural validity. The model was also assumed accurate with CMIN/DF= 1.796, parsimony comparative fit index= 0.753 and parsimony normed fit index= 0.717.

The standardised factor loadings and error variance for the final model are included within Table 2. The correlation between ‘Pain’ and ‘Symptoms’ was estimated at 0.642 with significant covariances (Estimate= 4.104, standard error= 1.101 and composite reliability= 3.728, p<0.001). The correlation between ‘Pain’ and ‘Function’ was estimated at 0.668 with significant covariances (Estimate= 5.424, standard error= 1.300 and composite reliability= 4.171, p<0.001). The correlation between ‘Symptoms’ and ‘Function’ was estimated at 0.275 with significant covariances (Estimate= 2.818, standard error= 1.353 and composite reliability= 2.082, p=0.037).

**Table 2.** Standardised Factor Loadings of the TENDINS-A (n=79)

| TENDINS- A | Factor | Standardised Factor Loading | Error Variance |
| --- | --- | --- | --- |
| Item 11 | Pain | 0.643 | 0.587 |
| Item 3 | Pain | 0.605 | 0.634 |
| Item 4 | Pain | 0.627 | 0.607 |
| Item 7 | Symptoms | 0.741 | 0.451 |
| Item 10 | Symptoms | 0.855 | 0.269 |
| Item 13C | Function | 0.921 | 0.152 |
| Item 13D | Function | 0.975 | 0.049 |
| Item 13E | Function | 0.967 | 0.065 |

#### Internal consistency

For the 8-items assessed using CFA included in TENDINS-A theorised to measure ‘disability’, the internal consistency was 0.842, hence displaying sufficient internal consistency.

#### Composite reliability

For the 8-items assessed using CFA included in TENDINS-A theorised to measure ‘disability’, the composite reliability was 0.934, hence displaying sufficient composite reliability.

#### Convergent validity

For the 8-items assessed using CFA included in TENDINS-A theorised to measure ‘disability’, the AVE was 0.648, hence displaying sufficient convergent validity.

Thus, our final CFA model was capable of determining the contribution of different items in measuring ‘Pain’, ‘Symptoms’ and ‘Function’ and the TENDINS-A has sufficient structural validity. Consequently, the scores for the remaining items were amended to ensure the TENDINS-A retained a maximal score of 100 (40 units for ‘Pain’, 30 units for ‘symptoms’ and 30 units for ‘function’). Questions 6 and 8 were also excluded from CFA modelling being dichotomous (Yes/No) items but were retained within the complete TENDINS-A in the ‘symptoms’ domain. All subsequent analyses therefore relate to the revised TENDINS-A, presented in Appendix E.

### Distribution of data

Whilst data for the TENDINS-A (Kolomogorov-Smirnov test statistic=0.09, p=0.097) and the VISA-A (Kolomogorov-Smirnov test statistic= 0.06, p=0.200) were normally distributed, the FAOS were not (Kolomogorov-Smirnov test statistic= 0.12, p=0.019). The TENDINS-A data had a minimum score of 0 and a maximum score of 100. The mean (SD) of the TENDINS-A was 47.89 (24.71) points, the FAOS was 23.60 (18.63) percent and the VISA was 53.58 (23.96) points. The mean (SD) of the repeated TENDINS-A was 44.34 (24.16) points.

### Reliability

#### Intraclass correlation co-efficient

The 10-item TENDINS-A mixed-effect ICC for absolute agreement of a single measure was ICC= 0.930 (95% CI = 0.881 to 0.959), p= < 0.001.

#### Internal consistency

The Cronbach’s Alpha for the 10-items of the TENDINS-A was reported as α=0.808, representing excellent internal consistency.

#### Standard error of the measurement

The TENDINS-standard error of the measurement was calculated as 6.54 units, indicating a relatively small standard error of the measurement.

### Minimal detectable change

The MDC for the 10-item TENDINS-A was calculated as 12.36 (95% CI= 5.46 to 19.25) units of difference, representing 25.6% points of change from a mean TENDINS-A score of 47.89.

### Convergent validity

The TENDINS-A exhibited a moderate positive correlation with the FAOS (rho=0.598, 95%CI= 0.408 to 0.738, p<0.001), and a moderate, negative correlation with the VISA-A (r=-0.639, 95%CI= -0.764 to -0.467, p<0.001).

### Divergent validity

The TENDINS-A showed no evidence of a statistically significant association with age (p=0.426) or BMI (p=0.189).

### Comparison between sub-groups

Between-group differences (Table 3) were observed for self-reported MVPA most days, with those performing moderate to vigorous physical activity most days having a lower TENDINS-A score (p=0.002) and males reported a lower TENDINS-A score (p=0.044). No differences in TENDINS-A score were observed for those performing specific tendon-loading exercise versus those who did not (p=0.079), or whether participants were multi-lingual (p=0.397). As more males performed MVPA than females in our sample, we performed a *post hoc* linear regression model to assess whether group differences due to sex were a result of higher levels of MVPA in males than females. The model supported this idea, with no association between the TENDINS-A score and sex (p=0.061) after adjusting for MVPA [17.7 (95%CI= 6.46 to 28.97) unit increase in TENDINS-A score, p=0.002].

**Table 3.** Between group differences in TENDINS-A score.

|  |  | Mean (SD),<br>range | Mean<br>Difference<br>(SED) | Test statistic<br>(Independent t-<br>test) | p-value |
| --- | --- | --- | --- | --- | --- |
| Moderate to<br>vigorous<br>physical activity<br>most days | Yes (n=55) | 37.69 (23.08),<br>0-91 | -18.81 (5.91) | -3.18 | 0.002* |
|  | No (n=24) | 56.50 (26.54),<br>5-98 |  |  |  |
| Sex | Male (n=39) | 37.56 (23.27),<br>5-98 | -11.54 (5.63) | -2.048 | 0.044* |
|  | Female<br>(n=40) | 49.10 (26.64),<br>0-91 |  |  |  |
| Tendon-loading<br>exercise<br>performed most<br>days | Yes (n=43) | 38.79 (22.35),<br>5-86 | -10.13 (5.69) | -1.779 | 0.079 |
|  | No (n=36) | 48.92 (28.23),<br>0-98 |  |  |  |
| Multilingual | Yes (n=28) | 48.79 (23.57),<br>5-91 | 8.34 (5.97) | 1.396 | 0.167 |
|  | No (n=51) | 40.45 (26.32),<br>0-98 |  |  |  |

Abbreviations: SD= standard deviation; SED= standard error of difference; n= number; range= min to max
\* $p < 0.05$

## DISCUSSION

The present investigation evaluated the construct validity (consisting of structural validity and hypothesis testing), as well as reliability (consisting of internal consistency, test-retest reliability and measurement error) of the TENDINS-A. The 10-item TENDINS-A, proposed to provide an overall measure of ‘disability’, demonstrated sufficient construct validity for three factors (pain; symptoms; function) and reliability, supporting its use in clinical and research settings. Furthermore, with normally distributed scores, there was no evidence of floor or ceiling effects within the sample. The TENDINS-A is also likely to be meaningful to people with a lived experience of Achilles tendinopathy, as the research team included such individuals to inform its development.^17^

All continuous variables of the TENDINS-A, after the removal of item 12, 13A and 13B, were able to load onto one of three factors: pain, symptoms or function. The core health domain of ‘disability’ is defined as the “composite scores of a mix of patient-rated pain and disability due to the pain, usually relating to tendon-specific activities/tasks,” and included in tools such as the VISA-A.^6 15^ Thus, the TENDINS-A is proposed to measure the overall composite of disability, using scores from items related to ‘pain’, ‘symptoms’ and ‘function’.

The reliability of the TENDINS-A is excellent (ICC>0.90). We opted to assess test-retest reliability on a single day as tendinopathy pain and symptoms are known to fluctuate daily.^20^ The internal consistency of the TENDINS-A was also excellent (α>0.80), ensuring all items were being scored consistently. The terminology surrounding the MDC, minimal important change and minimal clinically important difference (MCID) is confusing and for the purposes of discussion we have included all available comparisons. We calculated the minimal detectable change (MDC=12.4 units) of the TENDINS-A using a distribution-based method. This is larger than the MDC for the VISA-A, which is reported as between 7.8 to 8.2.^34^ A study using an anchor-based method in the VISA-A reported the MCID as 14 units,^35^ which is comparable to our estimated measure. Another study using the VISA-A estimated the MCID to be 6.5 units, however this was estimated from a selective sample of 15 participants with insertional Achilles tendinopathy, which may also explain the smaller MCID.^36^ To characterize the magnitude of change on the TENDINS-A that is meaningful to individuals with Achilles tendinopathy, future longitudinal studies are needed to estimate the minimal clinically-important difference (MCID).

The TENDINS-A demonstrated moderate correlations to both the VISA-A and FAOS, which is expected given they are proposed to measure the same overall health domain of disability.^15^ We did not expect strong correlations to the VISA-A or FAOS given the limitations of the VISA-A,^10 11 14^ which we theorised would extend to the FAOS as it is also lacking content validity. We expected the TENDINS-A was likely to be superior as it adhered to the guidelines set out by COSMIN for the development of an outcome measure. The lack of a strong correlation can also result from the absence of a gold standard outcome measure for disability.

Alternatively, we theorised the TENDINS-A should not have been associated with baseline characteristics such as age or BMI, and this was confirmed by our analysis. This differs to the VISA-A, whose total score is significantly associated with age and BMI,^37^ reflecting a problem for previous studies using the VISA-A that did not adjust for age or BMI within the statistical analysis.

None of our specified sub-groups (sex, being multilingual, MVPA most days, tendon-loading exercise most days) showed a floor or ceiling effect. Performing MVPA most days was associated with a lower TENDINS-A score, as expected. This is most likely a consequence of the numerous health benefits of physical activity.^38^ Alternatively, the inactive group may undertake less tendon-specific loading exercise and aggravate the Achilles less. However, this seems unlikely, as performing tendon-loading activity was also assessed and no significant effect detected.

### Limitations

The sample for this study, whilst diverse in some aspects, was heavily reliant on participants living in Australia. While other PROMS for Achilles tendinopathy showed acceptable cross-cultural validity in other languages,^39 40^ future research on cross-cultural adaptation of the TENDINS-A is needed.

Self-report may have resulted in a misdiagnosis of Achilles tendinopathy. However in this study, we enhanced diagnostic accuracy by implementing a standardized pain map ^18^. A prior investigation demonstrated a 93% concordance between patient-reported Achilles tendinopathy when using a pain map and diagnoses made by a physician.^19^ These findings suggest that, in the majority of cases, self-reported Achilles tendinopathy aligns with the clinical diagnosis.

Finally, further details related to medical co-morbidities and medication usage^41^ would have allowed analysis of their influence on total score and would be recommended in future research trials.

## CONCLUSION

Our evaluation of the revised 10-item TENDINS-A has determined it has adequate construct validity and reliability. These findings, and previously established content validity, ensure that the TENDINS-A can be recommended for immediate use in both research and clinical practice, being the preferred tool over the VISA-A and FAOS to assess disability in individuals with Achilles tendinopathy.

## Data Availability

All data produced in the present study are available upon reasonable request to the authors

## Appendix A. Summary of participant characteristics (n=79)

| Variable | Variable (sub variable) | n (%) |
| --- | --- | --- |
| Sex | Male | 39 (49.4) |
|  | Female | 40 (50.1) |
|  | Intersex | 0 (0.0) |
| Geographical location | Australia | 63 (79.7) |
|  | Netherlands | 10 (12.7) |
|  | United Kingdom | 3 (3.8) |
|  | United States of America | 2 (2.5) |
|  | Italy | 1 (1.3) |
| Multilingual | Yes | 28 (35.4) |
|  | No | 51 (64.6) |
| English first language (for those persons multi-lingual) | Yes | 15 (53.6) |
|  | No | 13 (46.4) |
| Ethnicity | Australian | 47 (59.5) |
|  | European | 20 (25.3) |
|  | Asian | 2 (2.6) |
|  | Indian | 2 (2.5) |
|  | North American | 2 (3.6) |
|  | New Zealander | 1 (1.3) |
|  | Other | 4 (5.1) |
|  | Decline to answer | 1 (1.3) |
| Physical activity | MVPA most days | 55 (69.6) |
|  | No MVPA most days | 24 (30.4) |
| Education | Less than high school | 3 (3.8) |
|  | High school graduate | 10 (12.7) |
|  | Bachelor's degree | 34 (43.0) |
|  | Master's degree | 20 (25.3) |
|  | Doctoral degree | 7 (8.9) |
|  | Professional degree (JD, MD) | 5 (6.3) |
| Employment | Full time | 50 (63.3) |
|  | Part time | 15 (19.0) |
|  | Home maker | 1 (1.3) |
|  | Student | 7 (8.9) |
|  | Retired | 3 (3.8) |
|  | Other | 2 (2.5) |
| Annual Income | Less than 30,000 AUD | 4 (5.1) |
|  | 30,000 – 49,999 AUD | 3 (3.8) |
|  | 50,000 – 79,999 AUD | 7 (8.9) |
|  | 80,000 – 99,999 AUD | 9 (11.4) |
|  | 100,000- 149,999 AUD | 19 (24.1) |
|  | 150,000 – 199,999 AUD | 12 (15.2) |
|  | Greater than 200,000 AUD | 22 (27.8) |
|  | Declined to answer | 3 (3.8) |
| Activities aggravating pain and symptoms from the TENDINS-A | Walking (slow) | 12 (15.2) |
|  | Walking (fast) | 35 (44.3) |
|  | Walking up and down stairs | 34 (43.0) |
|  | Running up and down stairs | 37 (46.8) |
|  | Running slow (jogging) | 48 (60.8) |
|  | Running fast (sprinting) | 43 (54.4) |
|  | Hopping and jumping | 49 (62.0) |
|  | Rapidly changing direction while running | 35 (44.3) |
| Activities typically performed in everyday life from the TENDINS-A | Walking (slow) | 65 (82.2) |
|  | Walking (fast) | 60 (75.9) |
|  | Walking up and down stairs | 62 (78.4) |
|  | Running up and down stairs* | 20 (25.3) |
|  | Running slow (jogging)* | 36 (45.6) |
|  | Running fast (sprinting)* | 21 (26.6) |
|  | Hopping and jumping* | 24 (30.4) |
|  | Rapidly changing direction while running* | 21 (26.6) |

\*Regarded as tendon loading activities
Abbreviations: n= number; %= percentage; MVPA= moderate to vigorous physical activity; JD= Juris Doctor; MD= medical doctor; AUD= Australian Dollars; TENDINS-A= Tendinopathy Severity Assessment – Achilles.

## Appendix B. Communalities for Exploratory Factor Analysis

| **TENDINS- A** | **Initial Communality** |
| --- | --- |
| Item 3 | 0.329 |
| Item 4 | 0.433 |
| Item 6 | 0.274* |
| Item 7 | 0.517 |
| Item 8 | 0.297* |
| Item 10 | 0.582 |
| Item 11 | 0.453 |
| Item 12 | 0.179* |
| Item 13A | 0.761 |
| Item 13B | 0.890 |
| Item 13C | 0.887 |
| Item 13D | 0.932 |
| Item 13E | 0.923 |

*\*Initial communality did not exceed 0.3.*

## Appendix C. Scree plot for principal axis factoring analysis

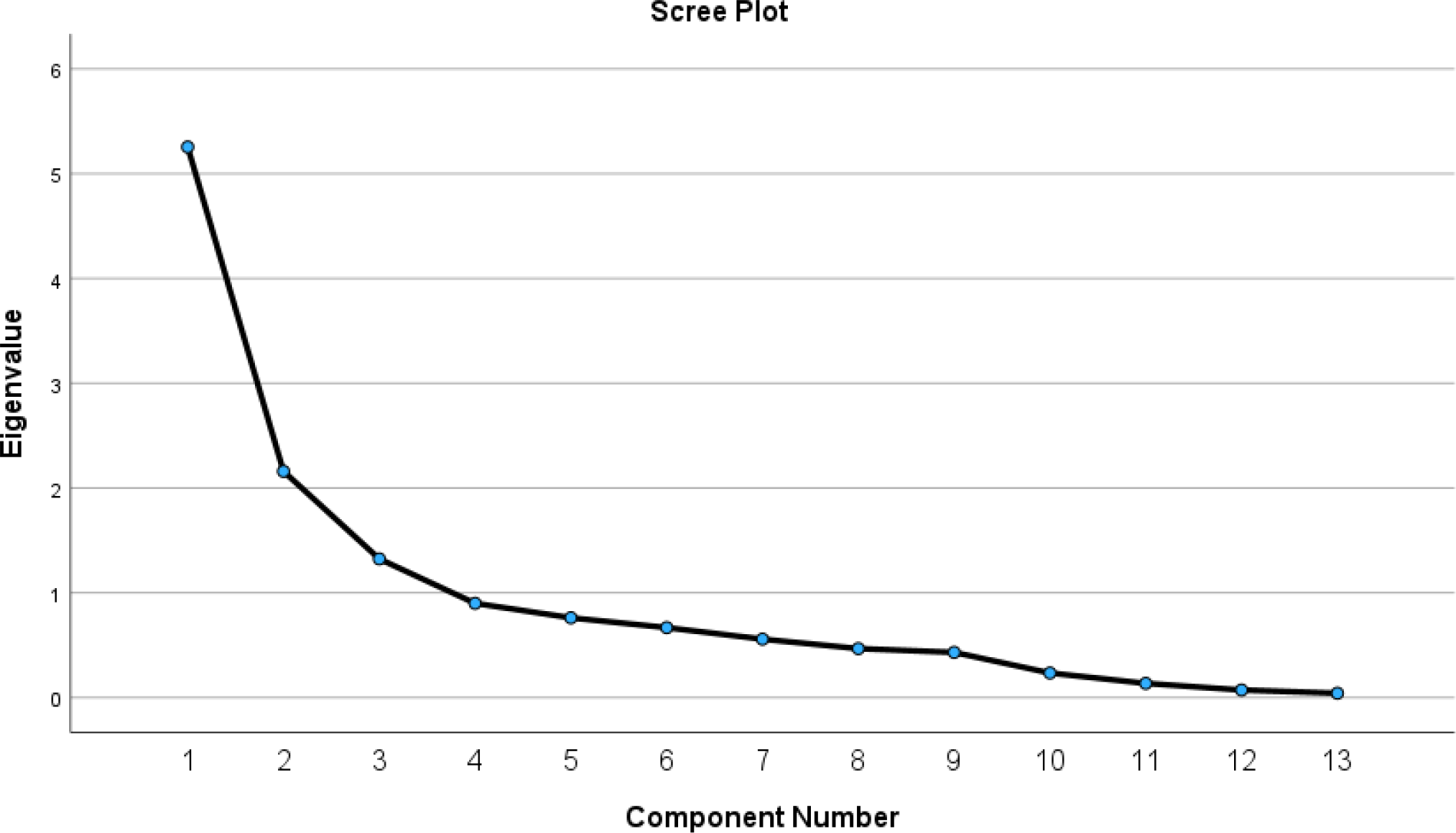

## Appendix D. Improvement in Model Fit following the exclusion of items 12, 13A and 13B

|  | **Model including all items** | **Model excluding items 12, 13A and 13B** |
| --- | --- | --- |
| **Comparative Fit Index** | 0.741 | 0.959 |
| **Tucker-Lewis Index** | 0.729 | 0.947 |
| **Root-mean-squared error of the approximation** | 0.189 | 0.101 |
| **Standardised Root Measure Square** | 0.1420 | 0.068 |

## Appendix E. TENDINopathy Severity assessment – Achilles (TENDINS-A)

This survey will ask you a number of questions about your Achilles tendon pain and symptoms. For the purposes of this survey, aggravating activities mean any activities that result in pain or symptoms, during or after their completion. For example, if you have pain during running it would be aggravating. Similarly, if you are stiff/tight/restricted the morning following fast walking this would also be aggravating. Your responses to question one and two will also be used as anchor points for the rest of the survey so please refer back to them as needed.

## UNSCORED SECTION

A. Please select as many of the following, if any, activities aggravate your Achilles tendon pain or symptoms?

For example, if you get pain running, but not walking you would select running but not walking. [Not scored]

□ Walking (slow)
□ Walking (fast)
□ Walking up and down stairs
□ Running up and down stairs
□ Running slow (jogging)
□ Running fast (sprinting)
□ Hopping and jumping
□ Rapidly changing direction while running
□ Other

B. Please select as many of the following activities you typically perform as a part of everyday life? For example, if you usually walk or jog but do not perform sprinting you would select walking and jogging but not sprinting. [Not scored]

□ Walking (slow)
□ Walking (fast)
□ Walking up and down stairs
□ Running up and down stairs
□ Running slow (jogging)
□ Running fast (sprinting)
□ Hopping and jumping
□ Rapidly changing direction while running
□ Other

C. If you perform the most aggravating activity from QUESTION (A) for your Achilles region PAIN (not stiffness) please record the following timepoint when your PAIN will be its worst.

□ Beginning or during the aggravating activity
□ Within two hours of stopping the aggravating activity
□ Within two to six hours of stopping the aggravating activity
□ Upon waking up the day following the aggravating activity

D. If you perform the activities from QUESTION (A) that aggravate your Achilles region STIFFNESS / TIGHTNESS / RESTRICTION please record the following timepoint when your STIFFNESS / TIGHTNESS/ RESTRICTION will be its worst.

□ Beginning or during the aggravating activity
□ Within two hours of stopping the aggravating activity
□ Within two to six hours of stopping the aggravating activity
□ Upon waking up the day following the aggravating activity

E. If you were currently made to perform the most painful/ symptom aggravating activity you recorded as being able to perform within everyday life within QUESTION (B) how long could you perform the activity before having to cease it due to your Achilles region pain/ symptoms?

□ I would not have to cease my activity due to Achilles region pain/ symptoms
□ Greater than 30 minutes
□ Between 15 to 30 minutes
□ Less than 15 minutes

## SCORED SECTION

### PAIN

1. How much have you had to change your current level of aggravating activities from QUESTION (A) in the unscored section since the onset of your Achilles region pain and symptoms?

□ I am currently performing my previous physical activity at a higher level than before my Achilles pain/symptoms [0]
□ I am currently performing my previous physical activity at the same level as before my Achilles pain/symptoms [0]
□ I am currently performing my previous physical activity at a lower level than before my Achilles pain/symptoms [10]
□ I am currently no longer able to perform any previous physical activity due to my Achilles pain/symptoms [20]

**In this entire section we are interested in the PAIN, and not other symptoms such as stiffness/tightness/restriction, you get within the Achilles region**.

2. When you are performing aggravating activities, does your Achilles region pain:

□ Improve (warm-up) [0]
□ Improve (warm-up) and then return while still performing your aggravated activity [3]
□ Stay the same [6]
□ Does not improve and gets worse the longer you perform the aggravating activity [10]

3. Does your Achilles region get painful once you have cooled down or rested following completion of the aggravating activities from QUESTION (A) in the unscored section?

□ No, I do not have any pain after I have cooled down or rested. [0]
□ Yes, I have pain after I have cooled down or rested and it lasts less than 15 minutes [3]
□ Yes, I have pain after I have cooled down or rested and it lasts between 15 to 30 minutes [6]
□ Yes, I have pain after I have cooled down or rested and it lasts greater than 30 minutes [10]

**PAIN SCORE:______ /40**

## SYMPTOMS

**In this entire section we are interested in the STIFFNESS/TIGHTNESS/RESTRICTION, and not pain, you get within the Achilles region**.

4. Do you typically have stiffness/tightness/restriction in your Achilles region upon getting out of bed after waking from sleep?

□ Yes [5]
□ No [0]

5. On average over the previous week, how long does your stiffness/tightness/restriction last upon waking up?

□ Less than 5 minutes [0]
□ 5-10 minutes [3]
□ 11-20 minutes [6]
□ Longer than 20 minutes [10]

6. Is your Achilles region stiffness/tightness/restriction upon waking worse the day after you perform the aggravating activities from QUESTION (A) in the unscored section?

□ Yes [5]
□ No [0]

7. After sitting for a prolonged period of time (two or more hours), if you then had to perform the aggravating activities from QUESTION (A) in the unscored section, how long would your Achilles take to loosen-up/warm-up?

□ Less than 5 minutes [0]
□ 5-10 minutes [3]
□ 11-20 minutes [6]
□ Longer than 20 minutes [10]

**SYMPTOMS SCORE: /30**

## PHYSICAL FUNCTION

### These movements relate to the questions below

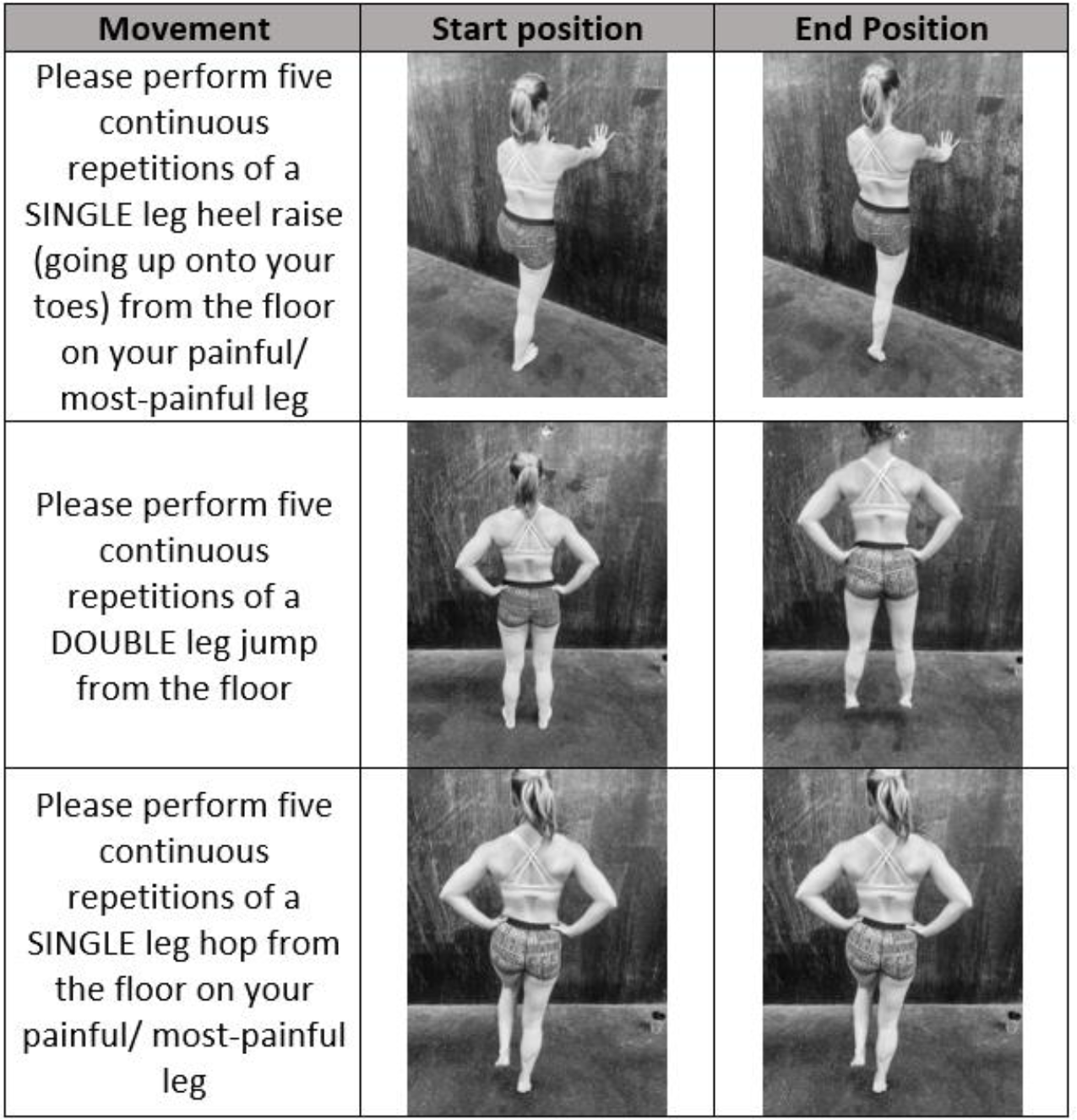

**For the following questions, please report your pain on a scale from 0 to 10, where zero is no pain and ten is the worst pain imaginable. If you are unable to complete the activity, leave the question blank. Images are provided to assist with positioning**.

8. When standing upright with knee fully straight, please perform five continuous repetitions of a SINGLE leg heel raise (going up onto your toes) from the floor on your painful/ most-painful leg and report the severity of your pain [0-10]

9. When standing upright with knees fully straight, please perform five continuous repetitions of a DOUBLE leg jump from the floor and report the severity of your pain [0-10]

10. When standing upright with knee fully straight, please perform five continuous repetitions of a SINGLE leg hop from the floor on your painful/ most-painful leg and report the severity of your pain [0-10]

**PHYSICAL FUNCTION SCORE:____ /30**

**TOTAL SCORE: /100**

